# Clofazimine pharmacokinetics in novel rifampicin-resistant tuberculosis regimens: an analysis of the endTB and endTB-Q trials

**DOI:** 10.64898/2026.09.18.26363422

**Authors:** Bernard Ngara, Belén P Solans, Eunsol Yang, Pieter Van Brantegem, Lorenzo Guglielmetti, Francis Varaine, Maelenn Gouillou, Carole D. Mitnick, Allison N. LaHood, Michael L. Rich, Kwonjune J. Seung, Lubbe Wiesner, Loren Hans, Rina Swart, Amanzhan Abubakirov, Kanat Khazhidinov, Anel Belgozhanova, Stephane Mpinda, Sesomo Mohale, David Holtzman, Dante Vargas Vásquez, Fanny García Velarde, Sergio Mucching-Toscano, Annum Aftab, Mahnoor S. Arshad, Azka Ashraf, Luong V. Dinh, Hanh T.T. Nguyen, Ha T.T. Phan, Sean Wasserman, Nelisiwe Ntuli, Radojka M. Savic, Helen McIlleron, Gustavo E. Velásquez, PandrTB Study Group

## Abstract

**Introduction:** Rifampicin-resistant tuberculosis poses a significant threat worldwide. Clofazimine is considered important for the construction of effective individualized multidrug regimens to treat rifampicin-resistant tuberculosis. Our goal was to characterize clofazimine pharmacokinetics in novel combination treatment regimens and to identify participant characteristics associated with variations in drug exposure.

**Methods:** Clofazimine pharmacokinetic data were obtained from adults and adolescents enrolled in the PandrTB pharmacokinetic sub-study of the endTB and endTB-Q Phase 3 randomized controlled therapeutic trials for rifampicin-resistant tuberculosis. Plasma concentrations were analyzed using nonlinear mixed-effects modeling to quantify clofazimine exposure and explore the impact of relevant covariates.

**Results:** 100 participants from six countries with high burdens of rifampicin-resistant tuberculosis contributed pharmacokinetic data. Their median age was 34 years, 32% were female, 18% were living with diabetes mellitus, and 18% were living with HIV. Clofazimine plasma concentration values were best described by a 2-compartment pharmacokinetic model with first-order absorption. Co-administration with delamanid and HIV co-infection resulted in a 37% increase and 20% decrease in clofazimine’s bioavailability, respectively. Diabetes was associated with a 43% decrease in clofazimine clearance. Clofazimine remained in the body for a median of 2.5 years (95^th^ percentile: 0.5 – 8) following 9 months of treatment.

**Conclusion:** Co-administration with delamanid, diabetes mellitus, and HIV were associated with variation in clofazimine exposure. We estimated that clofazimine remained present in the body for longer than two years in over half of participants following 9 months of treatment. Follow-up studies are recommended to confirm these associations before adjusting clofazimine dose or clinical care decisions.

## Introduction

Rifampicin-resistant (RR) tuberculosis (TB) remains a significant threat to TB treatment worldwide. In 2024, the World Health Organization (WHO) estimated that 390,000 people developed multidrug-resistant or rifampicin-resistant tuberculosis (MDR/RR-TB) globally [1]. The past decade has seen the rollout of safer, more effective therapies with shorter treatment durations [2]. There has been renewed interest in the use of clofazimine, an orally administered antimicrobial initially approved by the United States Food and Drug Administration (FDA) to treat leprosy [3], after regimens containing clofazimine showed potential to shorten the treatment duration of RR-TB safely and efficaciously [4,5]. In 2024 and 2025, WHO updated and consolidated recommendations that included multiple clofazimine-containing regimens [7,8]. A direct pharmacokinetic (PK) link between clofazimine exposure and its clinical use in these combination regimens has been lacking.

Clofazimine’s mechanism of action involves extensively binding to plasma proteins, accumulating in fatty tissues in a duration-dependent manner, and being sequestered in crystal-like drug inclusions inside macrophages [3,8–10]. Consequently, clofazimine has a long terminal half-life varying from 10 to 70 days in adults and has a large volume of distribution [11,12]. It is mostly excreted in its unchanged form [13] but is at least partly metabolized in the liver. Although the metabolism of clofazimine has not been clearly delineated, it involves several pathways, including oxidative, hydrolytic, and glucuronidation reactions. Key enzymes involved in these processes include CYP1A2, CYP3A4, and CYP3A5 for oxidative metabolism, and UGT1A1, UGT1A3, UGT1A4, UGT1A9, and UGT2B4 for glucuronidation [14]. Thus, potential pharmacokinetic (PK) drug-drug interactions may occur when clofazimine is co-administered with other drugs that induce or inhibit similar enzymes.

Earlier studies relied on two- or three-compartmental PK models with a lag-time or transit absorption and linear clearance. Variables such as sex, body fat, and diarrhea were found to be associated with clofazimine PK variability [12,15]. Here, we aimed to: (1) develop a population PK model, (2) investigate the effects of participant characteristics on PK variability, and (3) determine the duration of total elimination following the cessation of clofazimine in multidrug RR-TB regimens.

## Methods

### Source of data and population

Demographic, clinical, and plasma concentration data were obtained from participants enrolled in the PandrTB sub-study (NCT03827811) from 2019 to 2023 [16]. PandrTB was nested in the endTB (NCT02754765) [17] and endTB-Q (NCT03896685) [18] Phase 3 randomized clinical trials for RR-TB, sponsored by Médecins Sans Frontières. The endTB trial evaluated five 9-month all-oral regimens for treating fluoroquinolone (FQ)-susceptible RR-TB versus the local standard of care consistent with WHO guidelines. The endTB-Q trial was a randomized study for FQ-resistant RR-TB, evaluating a 4-drug oral regimen administered for 6 or 9 months depending on disease severity versus the local standard of care consistent with WHO guidelines. PandrTB study participants were adults and adolescents (≥15 years) recruited from seven sites in six countries that enrolled participants in endTB/endTB-Q: Kazakhstan, Lesotho, Pakistan, Peru, South Africa, and Vietnam. Plasma PK of the experimental arm medications was assayed. endTB, endTB-Q, and PandrTB were approved by the institutional or ethics review board that supervised each participating institution and clinical site. The University of Cape Town was the PandrTB institutional sponsor, overseen by the Human Research Ethics Committee at the University of Cape Town (HREC 739/2017). All participants provided written informed consent.

### Clofazimine dosing and pharmacokinetic sampling

Study participants received clofazimine for 9 months (endTB) or 6 to 9 months (endTB-Q) in combination with other anti-TB drugs. Clofazimine was administered at 100 milligrams (mg) daily across all treatment arms. PK sampling was performed on five different occasions: three during treatment (between 1 and 9 months post-randomization), and two post-treatment. At the first visit, participants were randomized to either semi-intensive or sparse PK sampling, while at subsequent visits, sampling was sparse only. During treatment, semi-intensive samples were obtained pre-dose and then 1.5, 3, 4.5, 6, 8, and 10 to 12 hours post-dose. Sparse samples were obtained pre-dose, and 8-10 or 24-28 hours post-dose, or, at 2-3 hours and 5-28 hours after the observed dose. After treatment, single-time-point samples were obtained at a convenient time up to 6 months post-treatment. On the PK sampling days, participants were provided food shortly before their morning doses of treatment, with the exact times of food intake, dose administration, and sampling recorded. The food-fat level was also estimated.

### Sample analysis

Clofazimine was analyzed with a validated liquid chromatography tandem mass spectrometry assay developed at the Division of Clinical Pharmacology, University of Cape Town [19].

### Population pharmacokinetic modeling and simulation

PK models with up to three compartments for disposition were tested [20]. The absorption process was assessed using first-order absorption with and without a lag time [21]. A series of transit compartment models were also evaluated for absorption [22]. Plasma concentrations below the lower limit of quantification occurred in fewer than 5% of all PK observations. These observations were excluded from analysis, consistent with convention [23]. Total body weight, fat mass, and fat-free mass were evaluated for inclusion in the base model to account for the effect of body size on clearance and volume of distribution parameters using allometric scaling [24]. Covariate modeling included co-administration of bedaquiline, delamanid, levofloxacin, linezolid, moxifloxacin, and pyrazinamide, along with additional covariates such as age, sex, HIV status, diabetes status, and food-fat level. The final model was used to calculate empirical values for the area under the plasma concentration-time curve over the final 24-hour dosing interval (AUC_0-24h_) using **Equation S1** (**Supplementary Appendix B**) and to assess the influence of various covariates on clofazimine exposure in study subjects. Simulations over several years after treatment cessation were performed in 1000 virtual individuals to investigate exposure and the duration needed to eliminate clofazimine from the body after treatment cessation.

### Software and statistical considerations

Model development, validation, and simulation were performed using nonlinear mixed-effects modeling using NONMEM version 7.5.1 [25], Perl-speaks-NONMEM version 5.4.0 [26], and R version 4.6.1 [27]. The PK model parameters were estimated using the first-order conditional estimation with interaction method. The best models were selected based on the change in objective function value at a 5% significance level, graphically by goodness-of-fit (GOF) plots, and by simulation-based diagnostics using visual predictive checks (VPCs). The stepwise covariate modeling [28] approach was used for covariate inclusion into the final model using a 5% and 1% level of significance for the forward inclusion and backward elimination steps, respectively. Bootstrapping was performed with 500 samples to determine 95% confidence intervals for the model parameters.

## Results

### Study participant characteristics

A total of 751 plasma PK concentrations of clofazimine were collected from 100 participants. Participants’ median age was 34 years, the median weight was 56 kg, and the median body mass index (BMI) was 20 kg/m². Comorbid HIV or diabetes mellitus each occurred in 18% of participants. Participants were assigned to regimens containing other anti-TB drugs as follows: 75% were concurrently receiving delamanid, 68% pyrazinamide, 57% bedaquiline, 46% levofloxacin, and 22% moxifloxacin (**Table 1)**.

**Table 1.** Characteristics of participants sampled for clofazimine pharmacokinetic analysis, stratified by treatment regimen. *B*, bedaquiline; *BMI*, body mass index; *C,* clofazimine; *D,* delamanid; *HIV,* human immunodeficiency virus; *L,* linezolid; *Lfx,* levofloxacin; *M,* moxifloxacin; *Z,* pyrazinamide.*, Some participants do not have HIV data.

| Variable | Study arm |  |  |  |  |
| --- | --- | --- | --- | --- | --- |
|  | BCLLfxZ<br>(n = 25) | DCLLfxZ<br>(n = 21) | DCMZ<br>(n = 22) | BDLC<br>(n = 32) | Total<br>(n = 100) |
| Age in years,<br>median (range) | 40 (21 – 62) | 32 (16 – 69) | 30 (15 – 56) | 39 (18 – 82) | 34 (15 – 82) |
| Female, n (%) | 11 (44) | 6 (29) | 6 (27) | 9 (28) | 32 (32) |
| Height in m,<br>median (range) | 1.64 (1.40 –<br>1.80) | 1.61 (1.50 –<br>1.83) | 1.68 (1.48 –<br>1.82) | 1.67 (1.40 –<br>1.84) | 1.65 (1.40 –<br>1.84) |
| Weight in kg,<br>median (range) | 57 (28 –82) | 58 (34 – 73) | 55 (45 – 83) | 55 (28 – 81) | 56 (28 – 83) |
| BMI in kg/m<sup>2</sup>,<br>median (range) | 21 (14 – 27) | 21 (13 – 27) | 20 (15 – 28) | 19 (14 – 28) | 20 (13 – 28) |
| Living with HIV, n<br>(%)* | 5 (20) | 2 (10) | 5 (23) | 0 | 12 (18) |
| Living with<br>diabetes mellitus, n<br>(%) | 3 (12) | 4 (19) | 2 (9) | 9 (28) | 18 (18) |

### Population pharmacokinetic modeling

A two-compartment PK model with a lag time and linear absorption and linear elimination best described clofazimine plasma concentrations using **Equations S2-S4** (**Supplementary Appendix B**), and the model schema is shown in **Figure 1**. The apparent clearance (*CL/F*) was estimated at 6 Lh^-1^ (RSE=7%), the apparent central volume of distribution (*V_c_ /F*) was estimated at 806 L (RSE=10%), and the first-order absorption rate (*K_a_*) was 0.39 Lh^-1^ (RSE=15%). The addition of an absorption lag of 0.2 (RSE=10%) hours significantly improved model fit, the apparent inter-compartmental clearance (*Q/F*) was 37 Lh^-1^ (RSE=7%), and the apparent peripheral volume of distribution (*V_p_/F*) was 13350 L (RSE=8%). Body weight outperformed fat mass or fat-free mass as a covariate on disposition parameters using allometric scaling and improved the base model fit. Concomitant use of delamanid increased clofazimine bioavailability (*F*) by 39% (RSE=22%), participants living with HIV had a 19% (RSE=47%) reduction in *F*, and participants with diabetes mellitus had a 45% (RSE=21%) reduction in *CL/F*. Inter-individual variability was 49% (RSE=9%) on *CL/F*, and 61% (RSE=9%) on *V_p_/F.* An inter-occasion variability parameter of 59% (RSE=10%) improved model fit when included in *F*.

**Figure 1.**
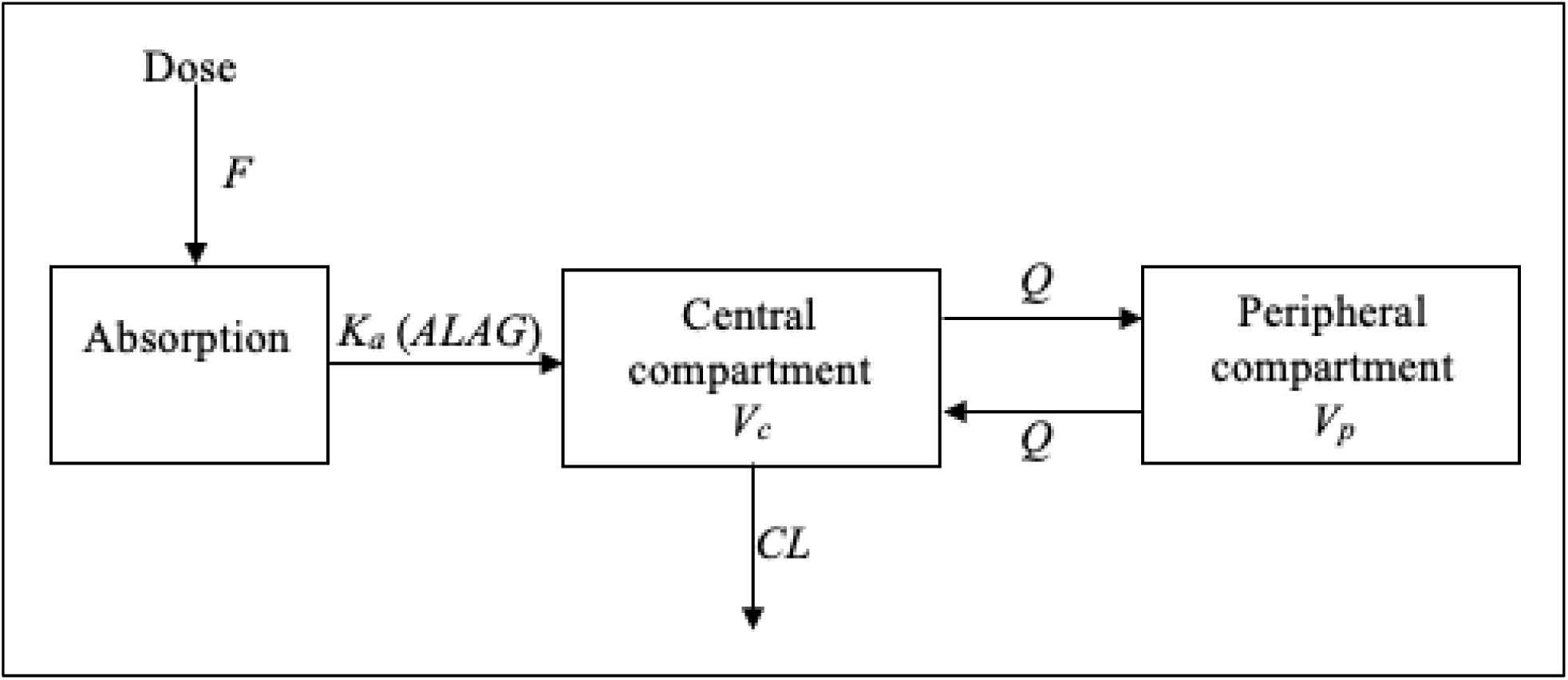
Structural model for clofazimine. The model describes total plasma concentrations. It consists of two compartments with linear absorption with a latency time, and linear elimination. *ALAG*, delay in the onset of absorption; *CL*, clearance; *F*, bioavailability; *K_a_*, absorption constant; *Q*, intercompartmental clearance; *V_c_*, volume of the central compartment; *V_p_*, volume of the peripheral compartment.

Residual error was estimated to be 14% (RSE=3%) using a proportional error model (**Table 2**). VPC and GOF plots show that the final model parameters adequately described clofazimine concentration data (**Figures 2 and 3**, respectively).

**Figure 2.**
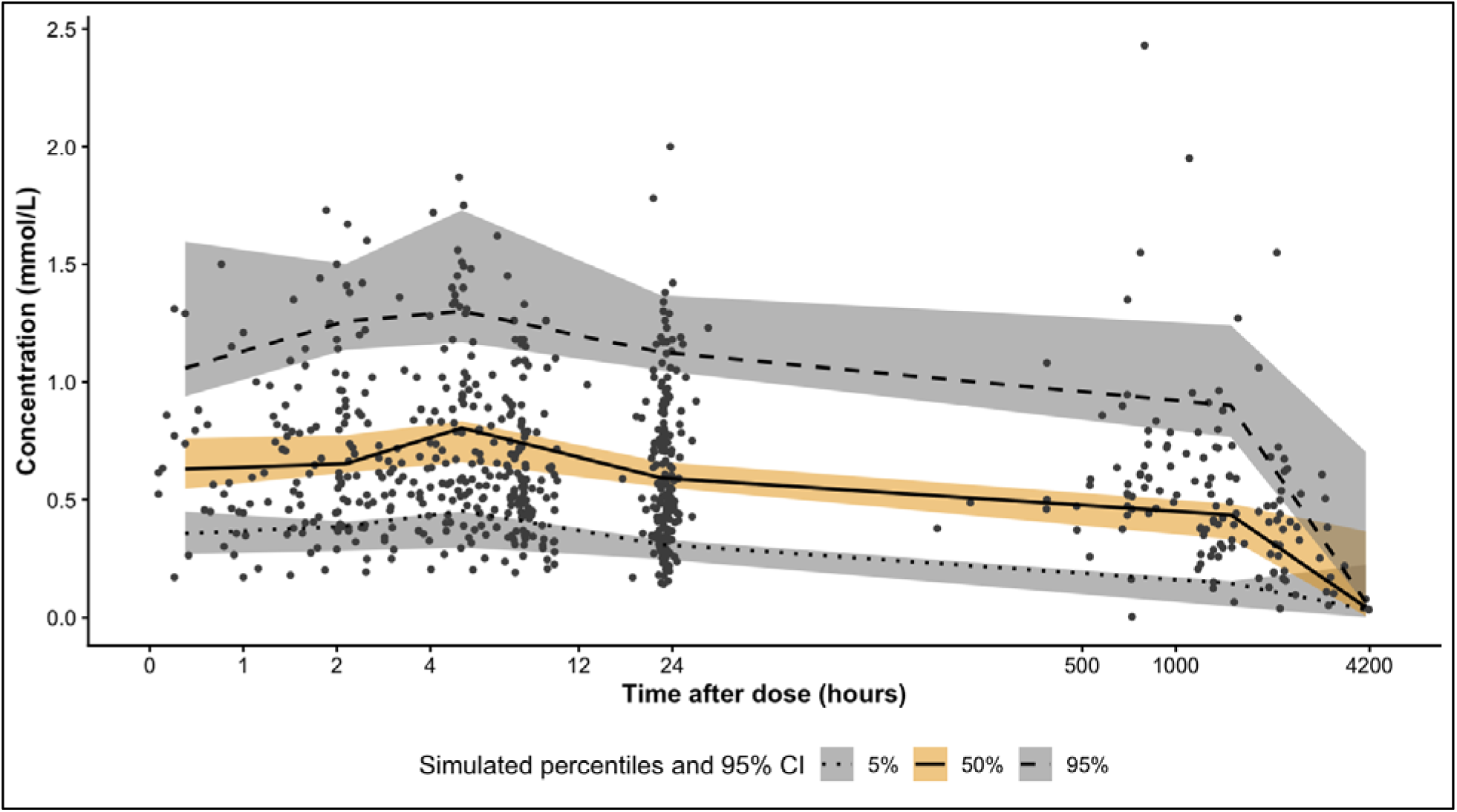
Prediction-corrected visual predictive check of the final clofazimine population pharmacokinetic model. The plot uses a log_10_ scale of time in hours after dose for the x-axis. Black dots represent observed individual clofazimine concentrations.

**Figure 3.**
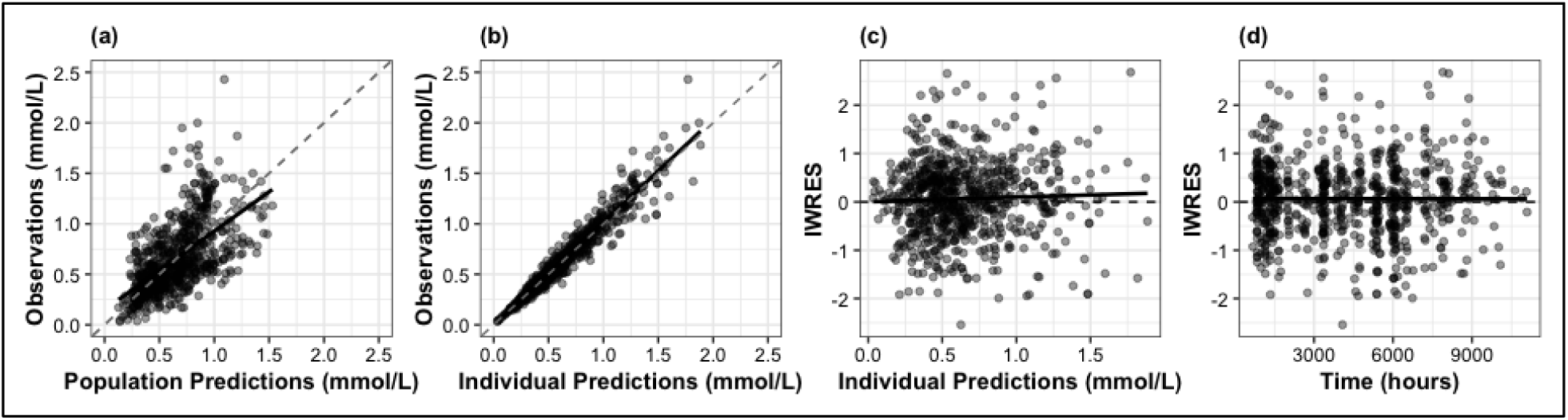
Goodness of fit plots of the final clofazimine population PK model. (**A**) observed versus population-predicted clofazimine concentration, (**B**) observed versu individual-predicted clofazimine concentration, (**C**) individual weighted residual errors versu individual-predicted clofazimine concentration, (**D**) individual weighted residual errors versu time after the first dose of clofazimine. Black dots represent individual values. Dashed lines represent identity lines. Solid lines represent the line of best fit for the scatter plot.

**Table 2.** Final clofazimine population pharmacokinetic model parameters. *\**, fixed parameter value; *ALAG*, delay in the onset of absorption; *CL/F*, apparent clearance; *CV*, coefficient of variation; *F*, bioavailability; *HIV,* human immunodeficiency virus; *IIV*, inter-individual variability; *IOV*, inter-occasion variability; *K_a_*, absorption constant; *Q/F*, intercompartmental clearance; *RSE,* relative standard error; *V_c_/F*, apparent volume of the central compartment; *V_p_/F*, apparent volume of the peripheral compartment. Body weight was included on *CL/F*, *Q/F*, *V_c_/F*, *V_p_/F* parameters using allometric scaling. Delamanid co-administration and HIV status were significant covariates adjusted for in the final model *F* parameter. Diabetes mellitus was a significant covariate adjusted for in the final model *CL/F* parameter.

| Parameter | Estimate (RSE %) | Bootstrap median (95% CI) |
| --- | --- | --- |
| Clearance ($CL/F$), $Lh^{-1}$ | 6 (7) | 6 (5 - 8) |
| Central volume of distribution ($V_c/F$), L | 806 (10) | 799 (523 - 1143) |
| First-order absorption rate ($K_a$), $Lh^{-1}$ | 0.39 (15) | 0.34 (0.21 - 0.51) |
| Inter-compartmental clearance ($Q/F$), $Lh^{-1}$ | 37 (7) | 40 (26 - 51) |
| Peripheral volume of distribution ($V_p/F$), L | 13350 (8) | 13809 (10643 - 18008) |
| Absorption lag time ($ALAG$), h | 0.2 (10) | 0.2 (0.1 - 0.29) |
| Bioavailability ($F$) | 1* | - |
| **Inter-individual variability (IIV)** |  |  |
| IIV on $CL/F$, CV% | 49 (9) | 23 (16 - 38) |
| IIV on $V_p/F$, CV% | 61 (9) | 36 (24 - 48) |
| **Inter-occasion variability (IOV)** |  |  |
| IOV on $F$, CV% | 59 (10) | 37 (20 - 55) |
| **Covariate effects** |  |  |
| Delamanid on $F$, % | 39 (22) | 40 (19 - 81) |
| Diabetes on $CL/F$, % | -45 (21) | -45 (-62 - -23) |
| HIV on $F$, % | -19 (47) | -18 (-28 - -8) |
| **Residual error** |  |  |
| Proportional error, % | 14 (3) | 14 (12 - 16) |

Model-predicted individual estimates for median AUC_0-24h_ were 24.1 mg.h/L (Interquartile Range: 18.7 – 32.8). Participants receiving delamanid had mean clofazimine exposure 36% higher compared to those not using delamanid (p < 0.05). Participants with diabetes mellitus had mean clofazimine exposure 79% higher than those without (p < 0.05). Participants living with HIV had mean clofazimine exposure 44% lower than those not living with HIV (p < 0.05) (**Figures 4a, 4b, and 4c**). Model simulations predicted that clofazimine remained in the body for a median period of 2.5 years (95^th^ percentile: 0.5 – 8 years) after treatment cessation (**Figure 4d**).

**Figure 4.**
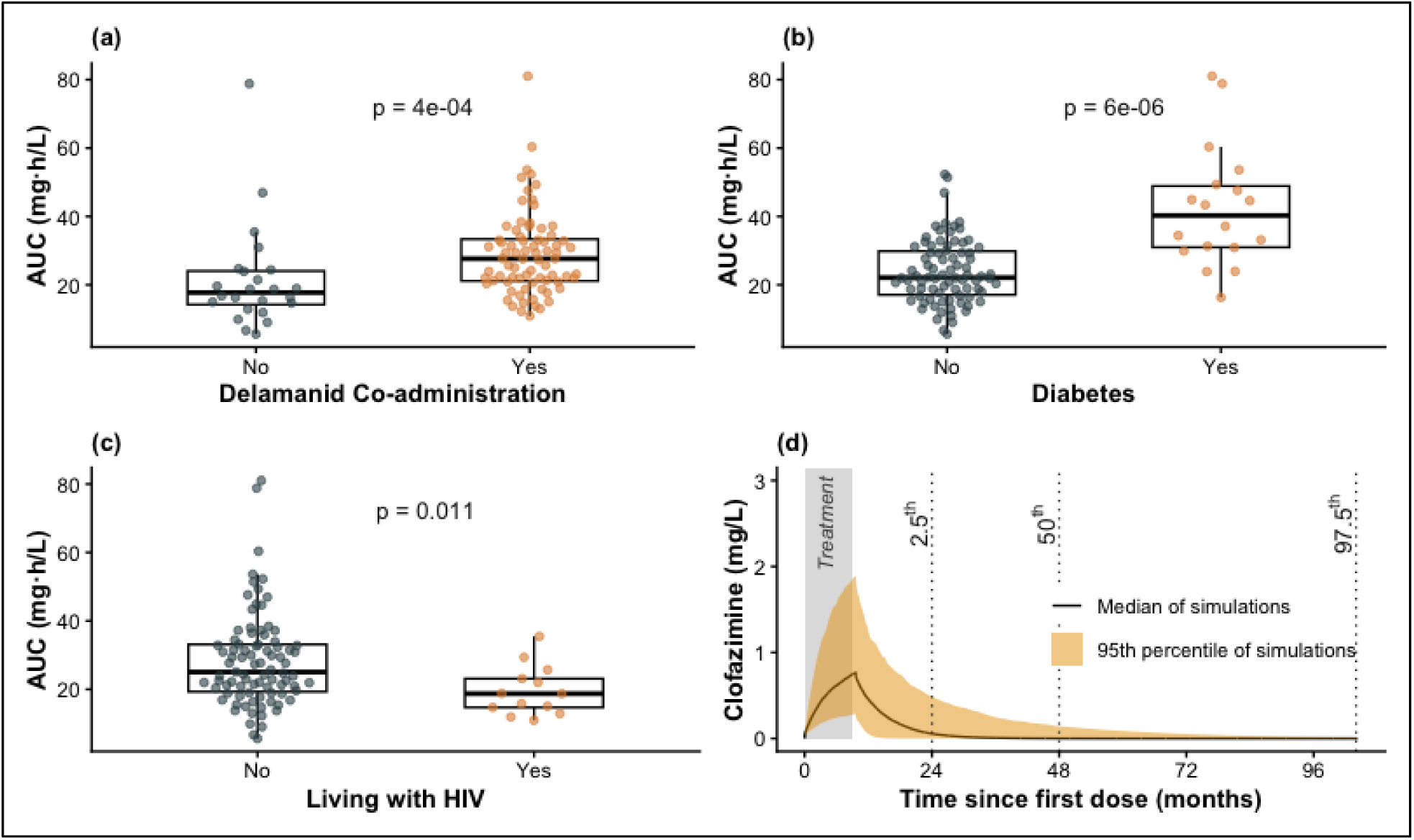
Box-and-whisker plots of the model-predicted individual estimates of the area under the concentration-time curve (AUC_0-24h_) for clofazimine following daily doses of clofazimine for 9 months. Plots are stratified by delamanid co-administration (a), a concurrent diagnosis of diabetes mellitus (b), and HIV status. The lower and upper hinges of each box correspond to the 25th and 75th percentiles, and the middle horizontal line of each box corresponds to the median. The whiskers extend from the hinge to the smallest or largest value no further than 1.5 times the interquartile range from the hinge. P-values were obtained using the Mann-Whitney U test. The results of simulations of clofazimine exposure, based on the final model, are illustrated in the bottom right panel (d).

## Discussion

To the best of our knowledge, this is the first study nested in clinical trials to evaluate clinical PK of clofazimine co-administered with three or four other anti-TB drugs in RR-TB patients across several continents. The study was novel in that PK data were collected up to six months after treatment cessation, which is helpful to enhance precise estimates of PK parameters like drug clearance of drugs like clofazimine with a longer half-life. Knowledge of the duration required for clofazimine to be cleared from the body is important in understanding the occurrence of clofazimine-related toxicity or the development of clofazimine-resistant strains if the infection is not totally cleared. The majority of research assessing clofazimine PK has relied solely on PK data gathered within 24 hours after the last dose [12,15,29], which may not accurately predict post-treatment exposure due to clofazimine’s previously reported long half-life ranging from 10 to 70 days [11,12].

This study found that the concurrent use of delamanid and the presence of diabetes mellitus in individuals were linked to higher clofazimine exposures. In vitro evidence linking diabetes and reduced CYP3A activity supports a mechanism for reduced clearance of clofazimine [30]. It is unclear whether co-administration with delamanid affects clofazimine clearance through specific pathways. That they may be potential confounders could not be addressed in this work and requires further investigation.

People with HIV having reduced bioavailability to clofazimine could be linked to antiretroviral therapy, HIV itself, or its effects on body composition. Diarrhea remains one of the most common side effects observed in antiretroviral therapy [31]. Clofazimine is a highly erratic drug that requires a prolonged absorption lag time and slow transit to be absorbed. Diarrhea can lead to HIV-associated hypermotility, physically sweeping the drug out of the gastrointestinal tract before significant absorption can occur. Recent clinical and pharmacokinetic evidence shows that severe diarrhea in HIV-infected individuals can lead to a higher than sixfold reduction (>83%) in clofazimine’s systemic bioavailability [32].

Final PK model simulations indicate that clofazimine is expected to remain detectable in the body for a median of 2.5 years following 9 months of daily doses of 100 mg. Earlier studies indicate that clofazimine is extensively distributed to peripheral compartments, followed by gradual re-equilibration to the central compartment [11,12,15,33]. Clofazimine is linked to the development of dry, rough, itchy, or scaly skin within weeks of starting treatment, which typically resolves 6-12 months after stopping the medication. However, some individuals may retain signs of discoloration for about 4 years [33]. Thus, as for bedaquiline, the development of resistance following treatment interruption of, or completion of, clofazimine-containing regimens is a concern, and our data may support the design of future approaches to reduce such risks [34,35].

A possible limitation for this analysis is the lack of information about the distribution of the CYP3A5 rs776746 single-nucleotide polymorphism, which could be detrimental considering these factors are associated with altered clofazimine exposures [36]. In conclusion, clofazimine is extensively distributed in the body and requires a median of 2.5 years to be eliminated from the body. Our results suggest that individuals with diabetes mellitus and co-administration with delamanid are probably experiencing elevated clofazimine concentrations, whereas those living with HIV have lower exposures. Further research is recommended to fully understand the mechanisms of interaction and develop clear guidelines for treatment modifications of clofazimine based on the results of this research. The clofazimine PK model will be useful in future exposure-response to determine associations of clofazimine PK with the efficacy and safety outcomes from the endTB and endTB-Q trials.

## Data Availability

All data produced in the present study are available upon reasonable request to the authors

## Acknowledgments

Author contributions.

B.N. wrote the first draft manuscript; B.N., B.P.S., E.Y., P.V.B., L.G., M.G., C.D.M., H.M., and G.E.V. reviewed and edited manuscript; B.N., B.P.S., and R.M.S. performed the data analysis; B.N., B.P.S., R.M.S., H.M., and G.E.V. interpreted results; E.Y., P.V.B., and M.G. performed data management; L.G., F.V., C.D.M., H.M., and G.E.V. designed the study; L.G., F.V., M.G., C.D.M., A.N.L, M.L.R., K.J.S., A.Ab., K.K., A.B., S.Mp., S.Mo., D.H., D.V.V., F.G.V., S.M.T., A.Af., M.S.A., A.As., L.V.D., H.T.T.N., H.T.T.P., S.W., N.N., H.H., and G.E.V. implemented the parent trials; A.N.L., A.Ab., K.K., A.B., S.Mp., S.Mo., D.H., D.V.V., F.G.V., S.M.T., A.Af., M.S.A., A.As., L.V.D., H.T.T.N., H.T.T.P., S.W., N.N., H.H., and G.E.V. implemented the study; L.W., L.H., and R.S. performed data generation.

## References

1. Global Tuberculosis Report 2025 [Internet]. [cited 2026 July 27]. Available from: https://www.who.int/teams/global-programme-on-tuberculosis-and-lung-health/tb-reports/global-tuberculosis-report-2025

2. Ndjeka N, Schnippel K, Master I, et al. High treatment success rate for multidrug-resistant and extensively drug-resistant tuberculosis using a bedaquiline-containing treatment regimen. European Respiratory Journal [Internet]. European Respiratory Society; 2018 [cited 2024 Sept 10]; 52(6). Available from: https://erj.ersjournals.com/content/52/6/1801528

3. Cholo MC, Steel HC, Fourie PB, Germishuizen WA, Anderson R. Clofazimine: current status and future prospects. J Antimicrob Chemother. 2012; 67(2):290–298.

4. Stadler JAM, Maartens G, Meintjes G, Wasserman S. Clofazimine for the treatment of tuberculosis. Front Pharmacol. 2023; 14:1100488.

5. Van Deun A, Maug AKJ, Salim MAH, et al. Short, highly effective, and inexpensive standardized treatment of multidrug-resistant tuberculosis. Am J Respir Crit Care Med. 2010; 182(5):684–692.

6. Pontali E, Raviglione M. Updated treatment guidelines for drug-resistant TB: how safe are clofazimine-based regimens? IJTLD Open. 2024; 1(11):486–489.

7. Conradie F, Badat T, Poswa A, et al. A Pragmatic Trial of a 6-Month Strategy for Rifampicin-Resistant Tuberculosis. N Engl J Med. 2026; 394(24):2429–2439.

8. Swanson RV, Adamson J, Moodley C, et al. Pharmacokinetics and Pharmacodynamics of Clofazimine in a Mouse Model of Tuberculosis. Antimicrob Agents Chemother. 2015; 59(6):3042–3051.

9. Schaaf HS, Garcia-Prats AJ, McKenna L, Seddon JA. Challenges of using new and repurposed drugs for the treatment of multidrug-resistant tuberculosis in children. Expert Rev Clin Pharmacol. 2018; 11(3):233–244.

10. Nugraha RV, Yunivita V, Santoso P, Aarnoutse RE, Ruslami R. Clofazimine as a Treatment for Multidrug-Resistant Tuberculosis: A Review. Scientia Pharmaceutica. Multidisciplinary Digital Publishing Institute; 2021; 89(2):19.

11. Schaad-Lanyi Z, Dieterle W, Dubois JP, Theobald W, Vischer W. Pharmacokinetics of clofazimine in healthy volunteers. Int J Lepr Other Mycobact Dis. 1987; 55(1):9–15.

12. Abdelwahab MT, Wasserman S, Brust JCM, et al. Clofazimine pharmacokinetics in patients with TB: dosing implications. J Antimicrob Chemother. 2020; 75(11):3269–3277.

13. Holdiness MR. Clinical pharmacokinetics of clofazimine. A review. Clin Pharmacokinet. 1989; 16(2):74–85.

14. Howlader S, Kim M-J, Jony MR, et al. Characterization of Clofazimine Metabolism in Human Liver Microsomal Incubation In Vitro. Antimicrob Agents Chemother. 66(10):e00565–22.

15. Zhang CX, Conrad TM, Hermann D, et al. Clofazimine pharmacokinetics in HIV-infected adults with diarrhea: Implications of diarrheal disease on absorption of orally administered therapeutics. CPT: Pharmacometrics & Systems Pharmacology. 2024; 13(3):410–423.

16. Study Details | Pharmacometrics to Advance Novel Regimens for Drug-resistant Tuberculosis-PandrTB Tuberculosis | ClinicalTrials.gov [Internet]. [cited 2024 Sept 11]. Available from: https://clinicaltrials.gov/study/NCT03827811?cond=pandrtb&rank=1

17. Guglielmetti L, Ardizzoni E, Atger M, et al. Evaluating newly approved drugs for multidrug-resistant tuberculosis (endTB): study protocol for an adaptive, multi-country randomized controlled trial. Trials. 2021; 22:651.

18. Patil SB, Tamirat M, Khazhidinov K, et al. Evaluating newly approved drugs in combination regimens for multidrug-resistant tuberculosis with fluoroquinolone resistance (endTB-Q): study protocol for a multi-country randomized controlled trial. Trials. 2023; 24(1):773.

19. Ali AM, P Solans B, Hesseling AC, et al. Pharmacokinetics and cardiac safety of clofazimine in children with rifampicin-resistant tuberculosis. Antimicrob Agents Chemother. 2024; 68(1):e0079423.

20. Mould DR, Upton RN. Basic Concepts in Population Modeling, Simulation, and Model-Based Drug Development—Part 2: Introduction to Pharmacokinetic Modeling Methods. CPT Pharmacometrics Syst Pharmacol. 2013; 2(4):e38.

21. Nerella NG, Block LH, Noonan PK. The impact of lag time on the estimation of pharmacokinetic parameters. I. One-compartment open model. Pharm Res. 1993; 10(7):1031–1036.

22. Savic RM, Jonker DM, Kerbusch T, Karlsson MO. Implementation of a transit compartment model for describing drug absorption in pharmacokinetic studies. J Pharmacokinet Pharmacodyn. 2007; 34(5):711–726.

23. Beal SL. Ways to Fit a PK Model with Some Data Below the Quantification Limit. J Pharmacokinet Pharmacodyn. 2001; 28(5):481–504.

24. Anderson BJ, Holford NHG. Mechanism-based concepts of size and maturity in pharmacokinetics. Annu Rev Pharmacol Toxicol. 2008; 48:303–332.

25. NONMEM | Nonlinear Mixed Effects Modelling | ICON plc [Internet]. [cited 2024 Sept 11]. Available from: https://www.iconplc.com/solutions/technologies/nonmem

26. Lindbom L, Ribbing J, Jonsson EN. Perl-speaks-NONMEM (PsN)--a Perl module for NONMEM related programming. Comput Methods Programs Biomed. 2004; 75(2):85–94.

27. R: The R Project for Statistical Computing [Internet]. [cited 2024 Oct 1]. Available from: https://www.r-project.org/

28. Jonsson EN, Karlsson MO. Automated covariate model building within NONMEM. Pharm Res. 1998; 15(9):1463–1468.

29. Stemkens R, Lemson A, Koele SE, et al. A loading dose of clofazimine to rapidly achieve steady-state-like concentrations in patients with nontuberculous mycobacterial disease. Journal of Antimicrobial Chemotherapy. 2024; 79(12):3100–3108.

30. Dostalek M, Court MH, Yan B, Akhlaghi F. Significantly reduced cytochrome P450 3A4 expression and activity in liver from humans with diabetes mellitus. Br J Pharmacol. 2011; 163(5):937–947.

31. Gupta R, Ordonez RM, Koenig S. Global Impact of Antiretroviral Therapy-Associated Diarrhea. AIDS Patient Care STDS. 2012; 26(12):711–713.

32. Zhang CX, Conrad TM, Hermann D, et al. Clofazimine pharmacokinetics in HIV infected adults with diarrhea: Implications of diarrheal disease on absorption of orally administered therapeutics. CPT Pharmacometrics Syst Pharmacol. 2024; 13(3):410–423.

33. Moore VJ. A review of side-effects experienced by patients taking clofazimine. Leprosy Review [Internet]. 1983 [cited 2025 Jan 22]; 54(4). Available from: http://leprev.ilsl.br/pdfs/1983/v54n4/pdf/v54n4a08.pdf

34. Gopal M, Padayatchi N, Metcalfe JZ, O’Donnell MR. Systematic review of clofazimine for the treatment of drug-resistant tuberculosis. Int J Tuberc Lung Dis. 2013; 17(8):1001–1007.

35. Park S, Jung J, Kim J, Han SB, Ryoo S. Investigation of Clofazimine Resistance and Genetic Mutations in Drug-Resistant Mycobacterium tuberculosis Isolates. J Clin Med. 2022; 11(7):1927.

36. Silva K, Ngara B, Van Brantegem P, et al. CYP3A5 polymorphisms influence bedaquiline and clofazimine pharmacokinetics in clinical trial participants with fluoroquinolone-susceptible and fluoroquinolone-resistant MDR/RR-TB across six countries. World Conference on Lung Health 2025 of the International Union Against Tuberculosis and Lung Disease. November 18-21, 2025, Copenhagen, Denmark.;

