## Supplementary appendix for "Clofazimine pharmacokinetics in novel rifampicin-resistant tuberculosis regimens: an analysis of the endTB and endTB-Q trials"

**A. List of endTB trial, endTB-Q trial, and PandrTB pharmacokinetic sub-study collaborators and contributors (*and study sites*):**

- **Dana-Farber Cancer Institute, USA:** Lorenzo Trippa
- **Epicentre, France:** Elisabeth Baudin, Romain Chenu, Souna Garba, Daiane Tozzi, Francis-Jaudel Yuya-Septoh
- **Hanoi Lung Hospital, Vietnam (*study site*):** Nguyen Thi Mai Anh, Pham Thu Anh, Phung Thi Bich, Dang Thi Hai, Nguyen Phuong Hoang, Nguyen Van Khiem, Pham Thi Lan, Bui Thi Tuyet Mai, Pham Thanh Nam, Dong Van Nguyen, Nguyen Thi Nhien, Nguyen Thi Kieu Ninh, Lai Thi Oanh, Thuong Huu Pham, Do Bang Tam, Dinh Thi Thuy, Trịnh Minh Trang, Tran Bao Trung, Nguyen Dinh Tuyen, Nguyen Hai Yen
- **Harvard Medical School, USA:** Eva Chang, Julia Coit, Jacquelyn-My Do, Kelsey O’Brien, Enoma Okunbor, Elna Osso
- **Hospital Nacional Hipólito Unanue, Peru (*study site*):** Shirley Carranza, Diego Delgado, Cemir Galarza, Gladys Murga, Elizabeth Ramos
- **Hospital Nacional Sergio Bernales, Peru (*study site*):** Kelly Cantaro, Andrely Huerta, Jonathan Maldonado, Roberto Nuñez, Katherine Panduro, Ruth Rafael, Epifanio Sánchez Garavito
- **The Indus Hospital & Health Network, Pakistan (*study site*):** Amaan Abdul, Sara Abedin, Eraj Afreen, Sohail Ahmed, Ubaid Ahmed, Jinsar Ali Shah, Faisal Aslam, Zahida Bashir, Saba Fayyaz, Kiren Hafeez, Meherunissa Hamid, Basmah Hassan, Muzamil Hussain, Muhammad Irfan, Maria Omar, Shafiqua Parveen, Sharoon Sunail, Muhammad Farooq, Batool Mazhar, Sana Munir, Hebah Mushtaque, Naseem Salahuddin, Neelofer Sonia, Muhammad Yaseen, Muhammad Zubair, Fatima Zehra, Iqra Zulfiqar
- **Institute of Chest Diseases, Pakistan (*study site*):** Tazeen Abbasi, Ghulam Akbar Abro, Mushtaque Ahmed, Nazir Ahmed, Muhammad Hammad Ali, Nazakat Ali, Rao Danish, Ayesha Farhat, Sunaina Gill, Muhammad Hafeez, Ghulam Hussain, Habib Inayat, Sylvia Johnson, Farees Kamal, Adil Kamran, Sanjeet Kumar, Mahveer Maheshwari, Shahid Mamsa, Sumera Massey, Asif Mehmood, Aaisha Memon, Ayaz Ali Mirani, Shafaque Naz, Tanzeel Rafique Qureshi, Yash Roop Moazzam Sheikh, Muhammad Rafi Siddiqui, Bhom Singh, Mustajab Soomro
- **Institute of Tropical Medicine, Belgium:** Elisa Ardizzoni, Bouke Catherine de Jong, Wim Mulders, Leen Rigouts, Praharshinie Rupasinghe
- **Interactive Development and Research, Pakistan:** Saman Ahmed, Muhammad Adnan Alamgir, Ayaz Ali, Azhar Ali, Shahzaib Ahmed Ali, Sakhawat Ali, Sanam Altaf, Samreen Amjad, Sadia Ausim, Abdul Basit, Haroon Ezik, Hira Hussain, Sibtain Hyder, Shahzad Inayat, Aleeza Janmohammed, Komal Jaseem, Aqsa Jawed, Lamis Maniar, Uzma Khan, Pardeep Kumar, Susheel Kumar, Reema Latif, Sara Liaqat, Muhammad Ammar Nasser, Muhammad Shakeel, Shahzaib Shaikh, Anique Siddiqui, Jetmal Singh, Koushalia Sivan, Momal Taimoor, Nabeel Zia
- **Médecins Sans Frontières, Armenia:** Ohanna Kirakosyan
- **Médecins Sans Frontières, United Kingdom:** Catherine Berry
- **Médecins Sans Frontières Khayelitsha, South Africa (study site):** Neide Cossa, Vivian Cox, Stobdan Kalon, Christine Knel, Nopinky Matinse, Vuyokazi Mbanjwa, Axole Ndayi, Tasanya Nomaliso, Tabassum Rashid, Allan Taylor, Hamza van der Ross, Brenda Wright
- **Médecins Sans Frontières OCG, Switzerland:** Sylvine Coutisson, Gabriella Ferlazzo, Nathalie Lachenal
- **Médecins Sans Frontières OCP, France:** Melchior Atger, Sandra Nadia Baya, Marwa Bekhiet, Veronique Boissière, Roberta Caboclo, Marhaba Chaudhry, Sandrine Cloez, Sandra Collin, Céline Delifer, Shana Desmaisons, Vanessa Ducher, Catherine Hewison, Mohamed Ibrahim, Kristen Lebeau, Merry Mazmanian, Rada Mirzayeva, Mathilde Moreau, Monica Moschioni, Audrey Pâquet, Christophe Perrin, Laura Pichon, Jeanne Roussel, Martina Scotton
- **MedStar Health Research Institute, USA:** Matteo Cellamare
- **National Lung Hospital, Vietnam:** Nguyen Kim Cuong, Nguyen Van Hung, Dinh Thu Huong, Le Thi Nguyet, Do Thi Thu, Do Thu Thuong, Nguyen Thi Thuy, Dam Truyen Thanh Tung
- **National Scientific Center of Phthisiopulmonology, Kazakhstan (*study site*):** Sagit Bektassov, Elmira Berikova, Lyailya Chingissova
- **Partners In Health Kazakhstan, Kazakhstan:** Zhanelya Amanzholova, Nazerke Birimkulova, Nurgul Dyusebayeva, Banu Kassenova, Tatyana Lee, Azhar Magzumova, Gulnar Omarova, Assel Stambekova, Gulmira Tanatarova, Zinatdin Uaisov, Zhanel Zhantuarova, Gulnara Zhumakairova
- **Partners In Health, USA:** Nataliya Arlyapova, Joshua Bogus, Meredith Cain, Merida Carmona, Clare Flanagan, Sarah McAnaw, Alyssa Scharff, Sonya Soni, Megan Striplin
- **Partners In Health Lesotho, Botsabelo MDR-TB Hospital, Lesotho (*study site*):** Seyfu Abebe, Johnson Alakaye, Atang Bulane, Michael Custodio, Precious Hajison, Malefetsane Hetsa, Matlotliso Khesa, Mikanda Kwabisha Kunda, Makatleho Lethola, Talime Lebitsa, Mpho Mahamo, Joalane Makaka, Mokenyakenya Matoko, Polo Mohoang, Daniel Monyaesa, Patrick Nkundayirazo, Seabata Ntsibane, Lawrence Oyewusi, Sebakeng Phate, Mathemba Radebe (Retsepile Tlali), Moliehi Rakhetsi, Tello Ranoosi, Meseret Tamirat Asfaw, Mahooe Thokoana
- **Partners In Health Peru (Socios En Salud Sucursal Perú), Peru:** Ericka Alegre, Christian Aguilar, Jessica Avalos, Lourdes Armuto, Nadia Barreda, Maria Barreto, Stephany Cabrera, Roger I Calderón, Milagros Castro, Jelina Chavez, Laura Chavez, Haydee Cheje, Lesli Cori, Daniel Dávalos, Silvana de la Gala, Karen Delgadillo, Hannely Diaz, Ximena Flores, Jimmy Galarza, Dalicxa García, Miriam Gaspar, Roselena Godos, Pamela Gomez, Luz Gonzales, Lizsery Guerrero, Sadith Inga, Judith Jasaycucho, Leonid Lecca, Bruno Martel, Ana Pro Martinez, Raquel Molina, Raquel Mugruza, Hansel Mundaca, Merilyn Nuahan, Kevin Ore, Liliana Panduro, Olga Peña, Sara Perea, Cynthia Pinedo, Yhomay Ponce, Claudia Quinte, Maritza Quiñones, Alicia Ramos, Elmer Ramirez, Rosina Reynoso, Cynthia Reyes, Jessica Reyes, Vanessa Riccio, Bryan Robles, Karen Rojas, Jimena Ruiz, Oswaldo Sanabria, Maria Saravia, Susana Saravia, Liz Senador, Jose Soberon, Edith Soncco, Maribel Soto, Catherine Suarez, Milagros Suarez, Veronica Suarez, Erika Torpoco, Oscar Torres, Claudia Torrez, Gabriela Tunque, Aida Ugarte, Gissela Vaderrama, William Valdivia, Yesica Valdivia, Isabel Valverde, Cynthia Vargas, Janet Vargas, Elvia Vasquez, Juan Veliz, Dioselinda Villa, Sheyla Villafuerte, Geraldine Villar, Stephanie Villegas, Milagros Wong, Rosa Yataco
- **University College London, United Kingdom:** Ilaria Motta
- **Université de Montpellier, Institut de Recherche pour le Développement, INSERM, France:** Maryline Bonnet
- **University of Cape Town, South Africa:** Nicole De Vries, Sehaam Jaffer, Marian Mazanhanga, Marilyn Solomons
- **University of California, San Francisco, USA:** Ariana Austin, Payam Nahid, Patrick PJ Phillips, Pearl Sun
- **Vietnam National University, Vietnam:** Nhung Viet Nguyen

**B. Supplementary Equations:**

${AUC}_{0-24h}=\frac{Dose\times F_{1}}{CL}$	**𝐸𝑞. S1**

$\frac{{dA}_{1}}{dt}=-K_{a}\times A_{1}$	**𝐸𝑞. S2**

$\frac{{dA}_{2}}{dt}=K_{a}\times A_{1}-\frac{CL}{V_{c}}\times A_{2}-\frac{Q}{V_{c}}\times A_{2}+\frac{Q}{V_{p}}\times A_{3}$	**𝐸𝑞. S3**

$\frac{{dA}_{3}}{dt}=\frac{Q}{V_{c}}\times A_{2}-\frac{Q}{V_{p}}\times A_{3}$	**𝐸𝑞. S4**

**Whereby:**

*AUC_0-24h_*: Area under the plasma concentration-time curve over the last 24-h dosing interval

$A_{1}$: Amount of drug in the gastrointestinal tract absorbed into the body

$A_{2}$: Amount of drug in the central compartment

$A_{3}$: Amount of drug in the peripheral compartment

$CL$*:* Clearance

$F_{1}:$ Bioavailability

$K_{a}:$ First-order absorption rate of the drug

$Q$: Intercompartmental clearance

$V_{c}$: Volume of the central compartment

$V_{P}$: Volume of the peripheral compartment
